# Real world costs and barriers to the successful implementation of rectal artesunate as pre-referral treatment for severe malaria in Sub-Saharan Africa

**DOI:** 10.1101/2022.05.24.22275488

**Authors:** Mark Lambiris, Guy Ndongala, Richard Ssempala, Victor Balogun, Michael Musiitwa, Fred Kagwire, Oluseyi Olosunde, Emmanel Emedo, Sylvie Luketa, Moulaye Sangare, Valentina Buj, Giulia Delvento, Katya Galactionova, Jean Okitawutshu, Antoinette Tshefu, Elizabeth Omoluabi, Phyllis Awor, Aita Signorell, Manuel W. Hetzel, Tristan T. Lee, Nina C. Brunner, Nadja Cereghetti, Theodoor Visser, Harriet G. Napier, Christian Burri, Christian Lengeler

**Author notes:** Corresponding author: Dr Mark Lambiris, Swiss Tropical and Public Health Institute, Socinstrasse 57, 4002 Basel, Switzerland. Equal contribution (Christian Burri, Christian Lengeler).

## Abstract

**Background:** Rectal artesunate (RAS), an efficacious pre-referral treatment for severe malaria in children, was deployed at scale in Uganda, Nigeria and DR Congo. In addition to distributing RAS, implementation required additional investments in crucial but neglected components in the care for severe malaria. We examined the real-world costs and barriers to RAS implementation.

**Methods:** We collected primary data on baseline health system gaps and subsequent RAS implementation expenditures. We calculated the equivalent annual cost of RAS implementation per child under 5 at risk of severe malaria, from a health system perspective, separating neglected routine health system components from incremental RAS introduction costs.

**Findings:** The largest baseline gaps were irregular health worker supervisions, inadequate referral facility worker training, and inadequate malaria commodity supplies. Health worker training and behaviour change campaigns were the largest startup costs, while supervision and supply chain management accounted for most annual routine costs. The equivalent annual costs of preparing the health system for treating severe malaria with RAS were $2.31, $2.20 and 4.15 per child at risk in Uganda, Nigeria and DRC. The incremental costs of introducing RAS, net of routine neglected components, accounted for a minority at $0.72, $0.59 and $0.94.

**Interpretation:** While RAS has been touted as a cost-effective pre-referral treatment for severe malaria in children, its real-world potential is limited by weak and under-financed continuums of care. Scaling up RAS or other interventions relying on community healthcare providers only makes sense alongside additional, essential health system investments sustained over the long-term.

**Funding:** Unitaid

## Introduction

Of the estimated 400’000 annual malaria deaths, the majority are in children below the age of five years living in Sub-Saharan Africa.^1^ Without prompt access to treatment with parenteral artesunate followed by oral artemisinin-based combination therapy (ACT), an episode of severe malaria in children can rapidly lead to death.^2^ Such comprehensive treatment presumes good access to higher level healthcare facilities. Poor children living in remote, rural settings are obviously challenged in accessing treatment and more likely to die from severe malaria.^3,4^

Community Access to Rectal Artesunate for Malaria (CARAMAL) was an observational study accompanying the roll out of rectal artesunate (RAS), an efficacious pre-referral treatment for severe malaria^5^, in highly endemic and difficult to reach rural settings in Uganda, Nigeria and the Democratic Republic of the Congo (DRC) targeted to children under 5 (CU5), under real world conditions. RAS, a suppository, rapidly reduces parasite density and buys a sick child time to reach a referral health facility that can treat severe malaria appropriately. Prior to CARAMAL, one large randomised controlled trial found that RAS reduced severe malaria case fatality by 26% (relative risk 0.74; 95% CI [0.59-0.93]).^6^

RAS was delivered in rural communities via routine case management: community health workers (CHWs) trained on integrated community case management (iCCM),^7^ and peripheral healthcare facilities (PHC) with no inpatient capacity. Implementation relied on appropriate training of health workers, supervision and a regular supply of drugs.^8-10^ Since the successful treatment of severe malaria relies on a cascade of healthcare services from the community until post-referral treatment completion, the CARAMAL intervention funded both the introduction of RAS into community-level structures and some operational strengthening of existing routine systems along the continuum of care. This “health system strengthening” (HSS) included the training of referral facility workers on parenteral artesunate, supervisions and some key supply chain inputs.

While several studies evaluated the costs of delivering services via CHWs^8,11-13^ for a range of diseases, this is the first study to our knowledge, that empirically assessed the real world costs of introducing RAS at community-level, on a large scale, including strengthening certain key parts of the cascade of care for children with severe malaria. In addition, we estimated the incremental cost of introducing RAS alone into an established system without additional HSS needs. In doing so, we documented important health system gaps, strategies implemented to overcome these, and their costs. The present analyses aim to inform operational guidance and financial planning in the replication or scale-up^14^ of RAS as pre-referral treatment for severe malaria. The findings also provide economists and modellers with real-world parameter costs towards economic evaluations of comprehensive interventions for severe malaria.

## Methods

### Implementation settings

The three settings differed markedly in the incidence of severe febrile episodes and the distribution of community-based providers and referral health facilities (RHF) - see Table 1. An overview of the whole project can be found in [Lengeler Burri et al. manuscript in preparation].

**Table 1:** number of children under 5 per healthcare provider and rate of severe febrile illness, by CARAMAL country. Note: numbers drawn from CARAMAL patient surveillance system. For details, see Lengeler Burri et al. manuscript in preparation

|  | Uganda | Nigeria | DRC |
| --- | --- | --- | --- |
| CU5 per community-based provider | 46 | 284 | 690 |
| CU5 per RHF | 11,816 | 55,022 | 7,045 |
| Rate of severe febrile illness per 1000 CU5 per year | 14.9 | 5.3 | 16.9 |

The implementation of the CARAMAL project and the introduction of RAS took place between Q4 2018 and Q4 2020 in Uganda, Nigeria and DRC. The intervention was implemented by local ministries of health supported by UNICEF (which we refer to as “implementers” throughout).

### Scope of the evaluation

Implementation activities were costed under a health system perspective and covered costs of services that would be incurred by the Ministry of Health to prepare the system to manage severe malaria with RAS. The analysis therefore excludes treatment costs of severe malaria and household costs (for these see Lambiris et al, in preparation), but includes the country’s procurement of RAS and injectable artesunate.

“Full implementation costs” are composed of two parts and labelled as either “startup” costs or annual “health system strengthening” (HSS) costs. Startup activities were one-time activities designed to launch the project. HSS were routine activities underlying the functioning of the existing health system (e.g. iCCM). We refer to these as “system strengthening”, rather than merely “routine”, to highlight that they either fully took over the funding of routine activities or complemented funding of essential, but often neglected, activities that national or donor financing was hitherto insufficient to cover. HSS activities recurred annually. A year’s worth of HSS was calculated from total expenditure per activity in the second year, per unit of time (quarters or number of months covered) before being converted to an annual cost.

We present these as economic costs expressed in real 2019 USD. Economic costs included level of effort costed via per-diems, time spent travelling and vehicle use, as well as donated commodities such as RAS and injectable artesunate adjusted to include cost, insurance and freight,^15^ using Global Fund prices.^16^ Research activities were excluded. Costs due to COVID-19 (e.g. PPE) were also excluded.

In addition to full implementation costs, we calculated the incremental costs of introducing RAS into a highly functional health system. These “RAS-specific” costs included activities that were additional to the routine components of the health system. RAS-specific costs included the proportion of training time judged specific to RAS; the procurement and distribution of RAS; the cost proportion of the initial behavioural change campaign relevant to RAS and severe malaria; and any novel elements that supported the introduction and maintenance of RAS that would not have been introduced otherwise. Expert opinion (UNICEF staff) decided these RAS-specific proportions. We calculated the RAS-specific costs for startup activities and annual HSS activities, separately, and present them as shares of full implementation startup and HSS costs.

Finally, we calculated the equivalent annual cost per child under 5 (CU5) at risk of severe malaria by dividing total equivalent annual cost by the total number of CU5 in the implementation areas.^17^ Annualisation allows converting the one-time startup cost into ten, equal, annual discounted net present costs over the project lifetime. We can thus sum the resulting annual startup and annual recurring costs into a single annual cost figure: the equivalent annual cost. We obtained the equivalent annual cost by annualising startup costs over 10 years, a time horizon reflecting longevity of a community-based programme (for formula see Appendix A),^11^ before adding the annually recurring HSS cost. We used a discount rate of 3%.^18^

### Implementation components

Below we describe the state of the health system prior to the intervention and country-specific gaps that were funded by CARAMAL. A detailed account of baseline and intervention components, both HSS and startup, can be found in Supplementary Tables S1a, S1b and S1c. Information on the baseline state of the health system was obtained from a healthcare provider survey conducted in Q4 2018 and RHF rapid readiness assessments in Q4 2017 [Lengeler Burri et al manuscript in preparation]. Information on baseline supervisory and behaviour change campaign (BCC) activity, as well as funding gaps were obtained from interviews with implementers throughout the implementation period.

#### Training

At baseline, CHWs and PHCs had not been trained on RAS. The intervention therefore included training, adapted for RAS integration: iCCM for CHWs or IMCI (Integrated Management of Childhood Illness) for PHCs. This training split was most notable in Nigeria where the system was characterised by a mix of many CHWs (2700) and PHC workers (806). It was less notable in Uganda that relied on CHWs mostly (4755 CHWs vs 86 PHC workers) and DRC where CHWs and PHC workers were trained together (242 health workers combined). In addition to startup training, refresher training was implemented in year 2 and assumed to recur every second year, as per national guidelines.

In all three countries at baseline, RHF staff characterised their training on management of severe malaria as inadequate or having occurred five or more years ago. 77% of RHFs in Uganda and 54% in Nigeria did not have a medical doctor trained on severe malaria while 31% of RHFs in DRC reported inadequate training on injectable artesunate. To ensure adequate case management for referred cases, CARAMAL covered the full cost of training RHF workers on severe malaria, malaria diagnostics and the role of RAS in community-referred cases; and the provision of injectable artesunate where lacking,

#### Supervision

Supervision at multiple levels was a recurring feature of the health system at baseline. Although national guidelines mandated them regularly, implementers in all three countries reported that one-to-one supervisions of CHWs and PHC workers often did not occur due to scarce and unsystematic funding.

During CARAMAL, CHWs were meant to restock on RAS while they met their supervisor. Under such circumstances, the absence of supervision implied RAS stock-outs. CARAMAL therefore covered the full costs of routine supervision (per-diems and travel expenses for supervisors or CHWs). In Uganda, one-to-one supervisions were scheduled biannually prior to the project. To increase oversight and minimise commodity stockouts in the community, implementers in Uganda increased supervisory frequency to quarterly. While systematic supervisions were a challenge in all countries, increasing their frequency was not necessary in DRC and Nigeria where they were supposed to occur on a monthly basis.

#### Procurement and supply chain

CARAMAL distributed RAS through the existing supply chain system in each country. Prior to this, however, stockouts in malaria commodities were reported frequently. Injectable artesunate stockouts in RHF were reported in the last 12 months at 69% and 95% of facilities in DRC and Uganda, respectively. In addition, more than 85% of CHWs reported stockouts of ACTs in the last 12 months in DRC and Uganda.

All three countries were unable to meet injectable artesunate demand from their national budget, so its procurement and distribution was fully costed, regardless of whether the funding came from CARAMAL or via Global Fund co-funding. In DRC, particularly, ongoing costs reflected considerable efforts to replace quinine with injectable artesunate during CARAMAL.

Finally, while supervisions doubled up as opportunities to re-stock CHWs with RAS frequently, these proved insufficient in Uganda. CARAMAL, therefore, funded parish coordinators with per-diems to restock CHWs with RAS on a monthly basis.

#### Behaviour change campaign

In all countries, behaviour change and education campaigns were described by implementers as donor-driven, time-limited and without systematic funding. In order to generate demand and awareness for RAS, posters and leaflets were printed, CHWs conducted home visits, community dialogues were held, and radio messages were aired. Some of these activities continued until the end of the project to ensure ongoing community education and involvement. We assumed, therefore, annual recurring costs covering educational components on malaria prevention, identification of danger signs, severe malaria case management and early health care seeking behaviour.

#### Monitoring and evaluation (M&E)

Staff time for monitoring the progress of implementation and, where necessary, financial support to local M&E systems, was accounted for. The latter was particularly important in DRC, where large gaps were financed, relative to Uganda and Nigeria.

#### Other supportive interventions

Ad-hoc interventions needed to overcome setting-specific obstacles in implementation were funded. Most notably, microscopes and hemoglobinometers were purchased for DRC referral facilities.

### Affordability

We estimated a proxy for the affordability of integrating RAS and HSS by comparing the public health expenditure per capita (World Bank^19^) to the recorded (non-discounted, non-annualised) implementation expenditures per capita. To obtain the latter we divided implementation startup costs and annual HSS costs by the total population in the study area. We then computed the ratio of implementation expenditures per capita to public health expenditures per capita.

### Data

Expenditure data provided by UNICEF was annual, between Q4 2018 and Q4 2020, and separate for Uganda, Nigeria and DRC. UNICEF determined the format expenditure data would be transferred to the research team in accordance with their institutional obligations. These were divided into implementation activities, and further disaggregated into sub-activities for which a total expenditure was given by year. Additional items were added by the research team and completed via interviews with UNICEF staff (sample expenditure table in Supplementary Table S2). Interviews aimed at obtaining in-depth understanding of activities and their purpose, as well as whether these complemented existing processes or supplementary activities were added to the health system for the project. Where co-funding from external donors was reported in annual reports or interviews, we did our best to obtain costs for these. Specifically, these included donations of injectable artesunate, RAS or co-funding of iCCM monitoring and evaluation systems. Relevant quantities such as number of RAS capsules or health workers trained were obtained from implementer interviews or CARAMAL annual reports.

## Results

### Economic costs of RAS integration and annual HSS

#### Startup expenditures

Full startup costs in real 2019 USD were $613’304, $997’338 and $709’575 in Uganda, Nigeria and DRC, respectively. Annual HSS costs were $612’033, $301’554 and $540’601 in Uganda, Nigeria and DRC. We present programme component shares of full implementation costs in Figure 1, Panel A, separately for startup and annual HSS costs (see Tables S1a, b and c for activity lists). Startup investments in health worker training accounted for large shares of full startup costs in the three countries. Training costs accounted for a greater share of startup costs in Nigeria relative to Uganda and DRC (61.2% vs 32.6% and 19.1% of startup costs respectively). The difference was due to transport and per-diems paid to Federal MoH officials (24% of total training costs) and the separate training programmes for CHWs and PHC workers in Nigeria, resulting in two sets of fixed costs (Supplementary Table S1b). These differences carry through to the training cost per CHW or PHC worker, which we calculate in Supplementary Table S3.

**Figure 1:**
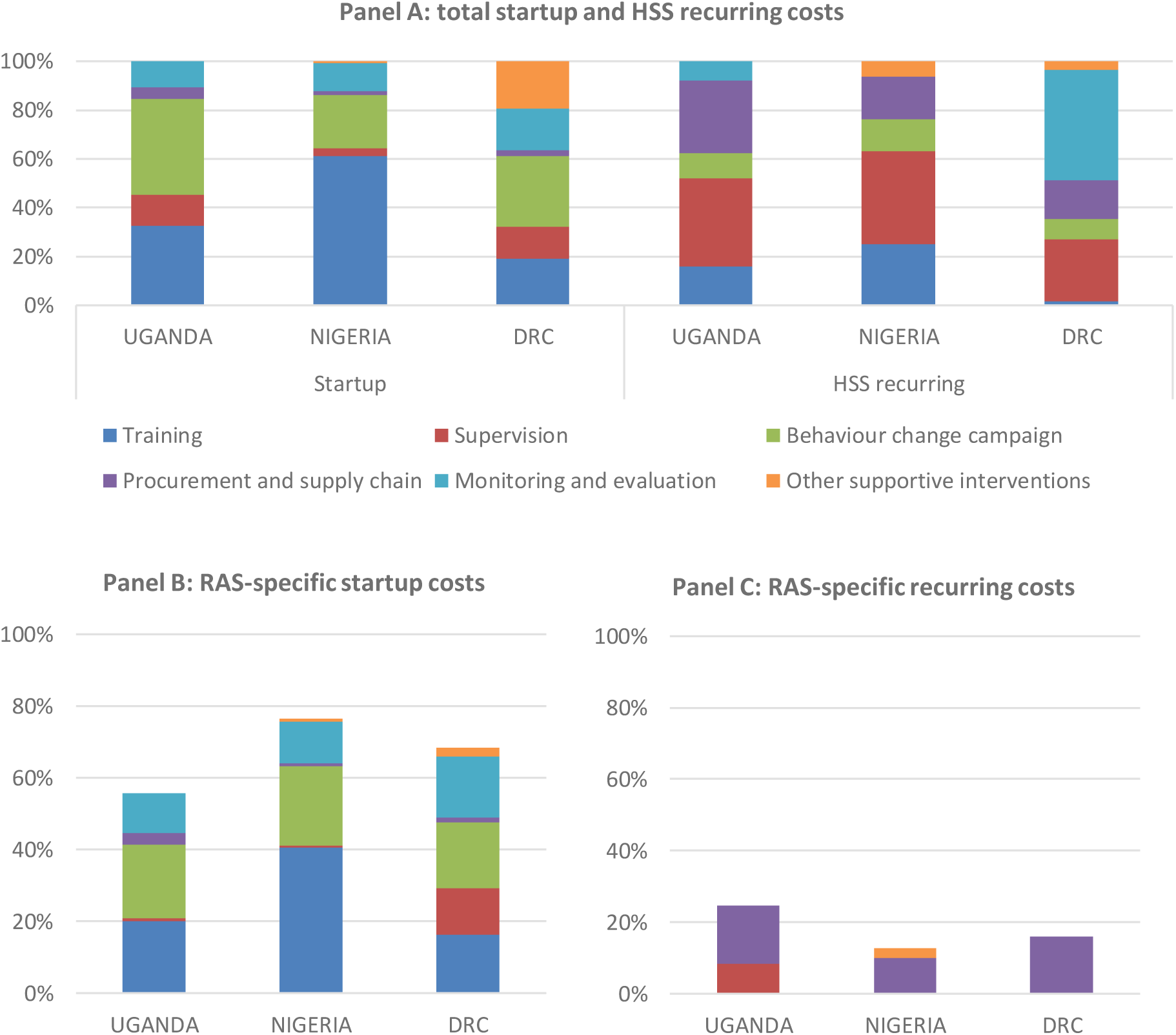
Total and incremental RAS-specific startup and recurring costs, by programme component. Note: Panel A shows the percentage of total intervention startup and HSS recurring costs that each programme component accounted for. Panels B and C show the RAS-specific proportion of the total presented in Panel A. The proportions are calculated from total costs in real 2019 USD. Total real 2019 USD startup costs were $613’304, $997’338 and $709’575 in Uganda, Nigeria and DRC, respectively. Annual HSS costs were $612’033, $301’554 and $540’601 in Uganda, Nigeria and DRC.

In addition to training, large investments were made in BCC. BCC startup activities accounted for 39.6% of startup costs in Uganda, 22.12% in Nigeria and 29.0% in DRC. Investments in other supportive startup activities were made in DRC (19.3% of startup costs), mainly towards strengthening the quality of care for severe febrile illness at RHFs. Startup supervision costs were small and covered either time-limited supervisions following training (DRC) or the printing of manuals and materials for supervisors.

#### Annual Health System Strengthening expenditures

Supervisions were the largest component of annual HSS costs. Recurring supervision costs amounted to 36.0% of annual HSS costs in Uganda, 37.9% in Nigeria and 25.1% in DRC (Figure 1, Panel A). We provide annual supervision unit costs per CHW for Uganda and Nigeria in Supplementary Table S3.

Annual supply chain costs were 30%, 17.7% and 15.8% of annual HSS costs in Uganda, Nigeria and DRC, respectively. Apart from the procurement of RAS in each country, sub-components varied. In addition to RAS, injectable artesunate was donated or procured and therefore costed annually. In Uganda, annual costs also included the monthly restocking of CHWs with RAS by parish coordinators (Supplementary Tables S1a, S1b and S1c), and commodity data collection. We present monthly unit costs per CHW for these supportive interventions in Supplementary Table S4.

### RAS-specific costs

Panels B and C of Figure 1 present the share of full startup and annual HSS costs (i.e. the share of costs presented in panel A), that are RAS-specific.

RAS-specific startup components in a functional and well-funded health system would cost 55.7%, 76.4% and 68.5% of actual startup costs in Uganda, Nigeria and DRC respectively (Panel B). Large initial health worker training costs (see Supplementary Tables S5 and S6) and investments in the BCC accounted for the majority of the cost of setting up RAS within the community-based health systems. In DRC, supplementary and time-limited MoH supervisions were conducted for several days every month, for three months after the completion of training. After this, routine and recurring supervisions took over.

The required investment to maintain RAS post-startup in a system that already funds its community-based programmes sustainably can be seen in Panel C. RAS-specific annual costs are a fraction of total annual HSS at 24.7%, 13% and 16% in Uganda, Nigeria and DRC. As expected, the bulk of these activity costs are the procurement and the distribution of RAS to CHWs and PHC workers. While these are similar shares in Nigeria and DRC, the share is higher in Uganda. As explained previously (Methods and Table S1a) implementers rolled out specific interventions to ensure RAS was systematically distributed to the large number of CHWs (nearly twice the number of CHWs in Nigeria and more than 100 times that in DRC). Jointly, these RAS-specific activities accounted for three quarters of the RAS-specific annual HSS costs.

### Economic costs per child under 5

The equivalent annual costs per CU5 at risk of severe malaria were $2.63 in Uganda, $2.20 in Nigeria and $4.15 in DRC (Figure 2). The costs for annual HSS made up the bulk of annual costs in all three project countries at 72.6% (Uganda), 72.7% (Nigeria) and 87.0% (DRC) of total annual implementations costs (Supplementary Table S7). The RAS-specific equivalent annual cost per CU5 is substantially lower in all three countries at $0.72 in Uganda, $0.59 in Nigeria and $0.94 in DRC or equivalently 31.4%, 26.9% and 22.6% of the full cost per child.

**Figure 2:**
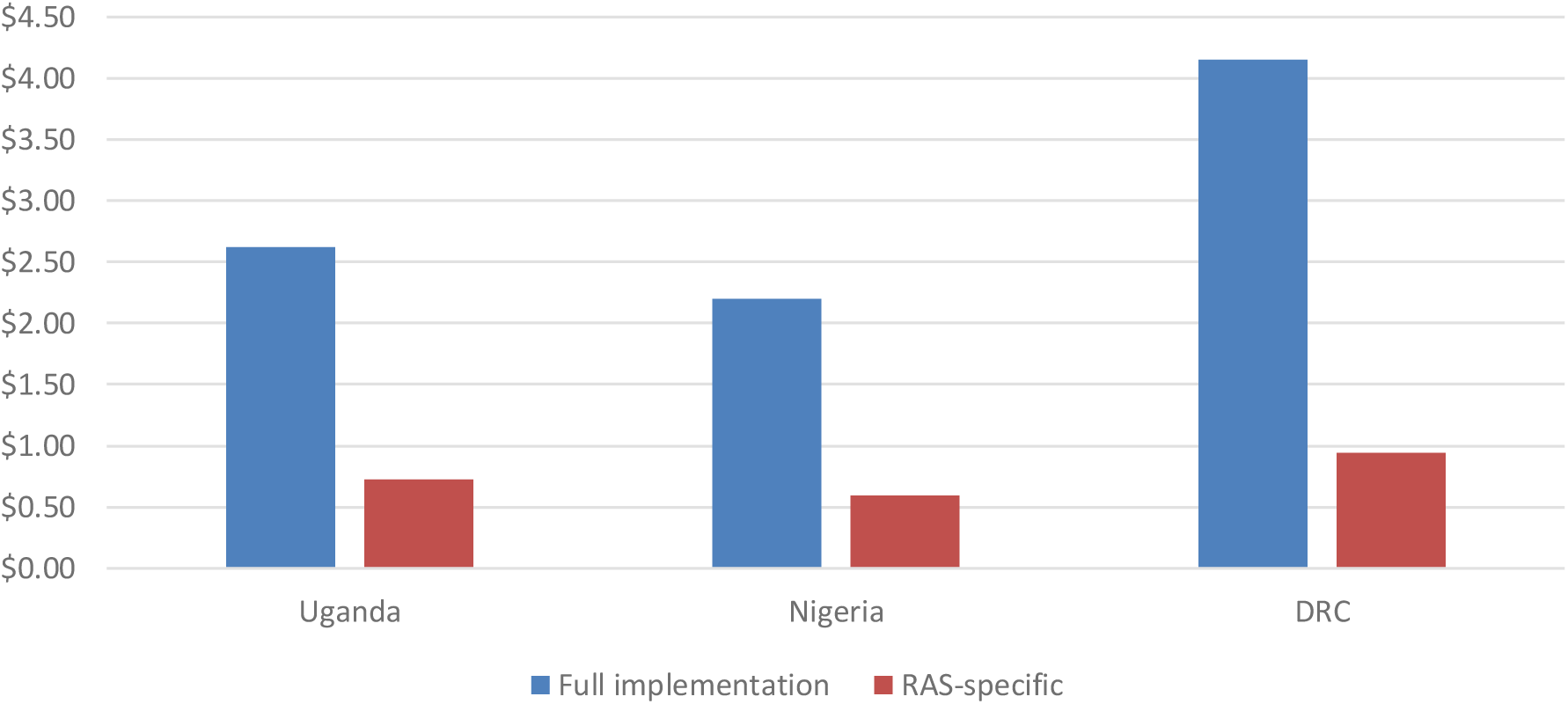
Full vs ‘RAS-specific’ equivalent annual cost of RAS implementation, per child under 5 years of age. Note: Costs are calculated as equivalent annual costs and in 2019 real USD. Startup costs were annualised over 10 years. The denominator is the total number of children in implementation areas, or otherwise: all children at risk of severe malaria. Number of CU5 covered by the implementation in Nigeria was calculated as the total number of CU5 in Adamawa state multiplied by the proportion of settlements in Adamawa covered by the iCCM programme, i.e. areas where the project was rolled-out (24.7%).

#### Affordability

Expenditures (financial non-annualised, non-discounted costs) during the start-up year (startup plus one year of HSS) amounted to 2.2%, 0.4% and 8.2% of the public health expenditure per capita in Uganda, Nigeria and DRC. For each year after that, HSS expenditures amounted to 1.1%, 0.1% and 4.1% of public health expenditures per capita. The substantially lower affordability in DRC is driven by a significantly lower per capita health expenditure ($18.52 per capita) compared to Uganda ($43.14) and Nigeria ($83.75).

## Discussion

CARAMAL introduced and monitored RAS in three distinct sub-Saharan African countries with high malaria burden, via community-level healthcare providers. Implementation leveraged pre-existing community-level health infrastructure to deliver RAS in remote settings where access to healthcare was poor. It further strengthened core system components in the cascade of care for severe malaria. Training, supervision, the supply chain, BCC, monitoring and evaluation and context-specific additional interventions were either strengthened operationally and financially, or adapted.

Using primary expenditure data and applying a health system perspective, we quantified startup activity costs and the annual health system strengthening costs required to prepare the health system for the effective management of suspected severe malaria cases in children under 5. The equivalent annual costs per child under five, annualised over 10 years, were $2.31 in Uganda, $2.20 in Nigeria and $4.15 in DRC, with the annual HSS component accounting for the largest share: 88.3%, 72.7% and 87.0% respectively.

These costs are high and reflect the low operational capacity and routine financing gaps in the continuum of care for severe malaria from community to tertiary care level. However, the vast gaps in annual HSS financing should also be strong cause of concern for other, new interventions that aim to be delivered via community-based healthcare systems. Without ensuring adequate funding and strengthened operational capacity the risk of failure remains high.

Due to CARAMAL’s focus on severe disease along the full continuum of care, comparing our estimates to costs of other malaria interventions might be misleading. A review of the costs of CHW programmes in low- and middle-income countries found only seven studies reporting these on malaria, with large heterogeneity in methods and scope.^8^ Among these, no studies focussed on severe malaria exclusively. No studies to our knowledge included the cost of training and supervising community-based providers, which included PHCs, beyond merely CHWs; or preparing referral-level facilities with training and commodity provision for treating severe malaria. Additionally, while we adopted a health system perspective here, other studies included patient-level costs, with large estimated indirect costs. While these societal perspectives are useful in their own right, they are beyond the present study’s scope. In spite of these differences, our estimate of the CHW unit cost of training, a more commonly-reported cost in other studies, lay in the broader range of other estimates in Sub-Saharan Africa.^11^ Finally, it is important to stress that the investment made to prepare the health system for the management of severe malaria would also benefit the treatment of other common diseases including diarrhoea and pneumonia, which are covered by iCCM.^20^ Subsequent cost-effectiveness analyses should include such benefits when trading them off against the large HSS costs.

Reported costs are not purely incremental. In theory, some included activity costs should already have been covered by the health system. In practice, however, activities such as supervisions were often not carried out prior to the intervention due to lack of funding. It was, however, not possible to ascertain the exact proportion of failed supervisions. In such cases, CARAMAL financed the full activity instead of just the incremental proportion. We followed suit in costing the activity in full so as not to underestimate financing requirements. This was particularly important for two reasons: (1) the lack of funding for supervision appears to be a systematic issue in iCCM and has been reported in other settings in Sub-Saharan Africa;^21-23^ and (2) strengthening gaps in supervisory activity operationally and financially also meant ensuring that RAS reached communities since supervisors often re-stocked CHWs directly.

While the above investments are necessary in preparing a functional continuum of care for severe malaria, they are likely insufficient to truly overcome access barriers and save the lives of those in the poorest and most remote locations.^24^ Sick children must complete referral – which was often not the case;^25^ and systematic post-referral treatment with artemisinin-based combination therapy must be guaranteed – which was also often not the case [Signorell et al. manuscript in preparation]. Only then could RAS realise its full potential and more young lives be saved. Until then, RAS is unlikely to be cost-effective as has previously been claimed under controlled conditions.^26^

In addition to full implementation costs, we estimated the incremental cost of introducing the commodity into the system, net of routine components. This “RAS-specific” cost was $0.72, $0.43 and $0.64 per child. These represented 35.3%, 19.5% and 14.7% of the full equivalent annual costs in Uganda, Nigeria and DRC. Obviously, it would be significantly less costly to introduce RAS into settings where iCCM is already adequately financed and functioning. The supply chain management of RAS was the costliest component; and particularly the distribution of RAS from supervisory health facilities to communities. In Uganda, the recruitment of parish coordinators to supply RAS and the institution of additional yearly supervisions increased both RAS availability but also the costs relative to Nigeria and DRC. Re-supplying could be eased in the future if the shelf life of RAS increased beyond the current 3 months. The latter seems plausible since guidelines were recently changed allowing for the shelf life of RAS to increase from 3 to 6 months (Medicines for Malaria Venture, private communication).

Finally, affordability of the intervention was substantially more favourable in Uganda and Nigeria than in DRC, where public health expenditures were the smallest. The startup year amounted to 2%, 0.4% and 8.2% of the public health expenditure per capita in Uganda, Nigeria and DRC and 0.9%, 0.1% and 4.1% for every subsequent year after that. The DRC numbers are concerning considering that donor-driven contributions in DRC have dropped from 43% to 35% of total public health expenditures per capita between 2016 and 2018 at a time when total health expenditures per capita in DRC have decreased by $2.^19^ More broadly, it remains a stark reality that many iCCM systems in Sub-Saharan Africa are largely dependent on donor funding.^27^ Our study confirms that partial financing cannot sustain complex community health systems. Unless donor funding streams are aligned, harmonised and sustained over the long-run it seems unlikely that health system gaps, access to treatment, and reductions in malaria mortality will resolve.

## Data Availability

The data may be made available post-publication on agreement of all contributing parties.

## Acknowledgements

We thank the health workers, local and national health authorities who provided their support; all UNICEF implementation teams in Uganda, Nigeria and the Democratic Republic of the Congo; our partners at the School of Public Health in Kinshasa (DRC), Akena Associates (Nigeria), and Makerere University School of Public Health (Uganda); and the colleagues of the local CHAI, UNICEF and WHO offices.

## Authors’ contributions

ML, CB and CL conceived the study. ML designed the methodology, led the formal analysis and the data collection process. GN, RS and VBa contributed to data collection, data curation and analysis. MM, FK, OO, EE, SL, MS and VBu provided the data. NC, VBu, HN and TV provided project and coordination support. ML wrote the first version of the manuscript. All authors contributed to reviewing and editing the manuscript. All authors approved the final draft of the manuscript.

